# POST-COVID-19 SYNDROME, INFLAMMATORY MARKERS AND SEX DIFFERENCES

**DOI:** 10.1101/2021.07.07.21260092

**Authors:** Meryam Maamar, Arancha Artime, Emilio Pariente, Patricia Fierro, Yolanda Ruiz, Silvia Gutiérrez, Roberto González, Elena Bustamante, Gabriel Pinedo, Blanca Rodríguez, Alvaro Peña, Miguel A Gómez, Celeste Urarte, Isabel Pérez-Pajares, Marian Tobalina, Carmen Secada, Sara Díaz-Salazar, Stefanie Pini, Carmen Ramos, José M Olmos, José L Hernández

## Abstract

**INTRODUCTION AND OBJECTIVE:** Post-COVID syndrome (PCS) is a poorly-known entity. Underlying low-grade inflammation (LGI) has been theorized as one of its pathophysiological mechanisms. We aimed to investigate a possible relationship between PCS and an increase in inflammation markers, in a sex-stratified analysis.

**PARTICIPANTS AND METHODS:** Mild cases of COVID-19 according to the WHO classification followed-up in a Primary Care Center, were included. We collected epidemiological data (age, sex, body mass index -BMI-, smoking, and comorbidities -Charlson index-), variables of the acute COVID-19 episode, and data at 3 months of follow-up (clinical manifestations and inflammatory markers). Serum C-reactive protein (CRP), neutrophil and lymphocyte counts, neutrophil/lymphocyte ratio (NLR), lactate dehydrogenase (LDH), ferritin, fibrinogen, and D-dimer levels were analyzed. Low-grade inflammation (LGI) was defined as serum CRP between >0.3 and <1.0 mg/dL. Five composite indices were built combining the upper ranges of 4 markers. Bivariate and multivariate analyses (logistic regression and general linear models) were performed, stratified by sex.

**RESULTS:** We analyzed 121 subjects with mild COVID-19 (56.2% women; mean age 46 years). The most common symptom in the acute episode was fever (60.3%), while it was fatigue in PCS (42.8%). Prevalence of PCS was 35.8% in women and 20.8% in men (p = 0.07).

In women, after controlling for age, BMI, smoking, and comorbidities, the D1, D3, and D4 indices were consistently associated with PCS, with ORs of 5.14 (95% CI, 1.6-16.4), 4.20 (95% CI, 1.3-13.3), and 4.12 (95% CI, 1.3-13.1), respectively; in patients with post-COVID anosmia and ageusia, neutrophils were significantly elevated (3.43±0.3 vs 2.58±0.1; p = 0.014, and 3.89±0.3 vs 2.59±0.1; p = 0.002,respectively), after adjusting for confounders.

In men, the D2 and D5 indices were associated with PCS, with adjusted ORs of 10.1 (95% CI, 1.2-85) and 17.5 (95% CI, 2-153), respectively. Furthermore, serum CRP in the LGI range was associated with PCS [adjusted OR=12.9 (95% CI, 1.3-121)], and in post-COVID persistent fatigue, the neutrophil count was significantly elevated (4.68±0.6 vs 3.37±0.1; p = 0.041), after controlling for confounders.

**CONCLUSIONS:** Consistent associations among PCS, anosmia, ageusia, and fatigue, with slight -but significant-elevated levels of inflammatory markers, have been observed. The neutrophil count was the most frequently involved marker. Sex-stratified analyses showed relevant differences between women and men concerning PCS and serum inflammatory markers.

## INTRODUCTION

Post-COVID-19 syndrome (PCS) includes a large number of long-term and recurrent symptoms experienced by a significant percentage of patients recovering from acute COVID-19. Fatigue, myalgia, dyspnea, anosmia, ageusia, autonomic dysregulation (manifested as orthostatic hypotension, tachycardia, thermoregulatory or gastrointestinal alterations), anxiety, depression, and cognitive problems were the main symptoms reported [1,2]. These clinical manifestations fluctuate over time [3], and can significantly affect the quality of life [4].

To date, little is known about this syndrome and there is no clear correlation between the severity of the acute viral episode and the subsequent appearance of PCS [5]. Risk factors for having severe acute COVID-19, such as age, male gender, or obesity, have not been fully related to the development of PCS [6]. It is still unknown if this syndrome is a final extension of the COVID-19 infection or a different entity. From a pathophysiological point of view, it has been suggested that a sustained viral replication, microbiome alterations, a dysregulated immune/inflammatory response, or a disorder similar to chronic fatigue syndrome (CFS), might be involved in the PCS development [6-8].

Of these, the hypothesis of the dysregulation of the immune system and an inflammatory state as the pathophysiological basis of PCS is generating increasing interest [7,9]. It is well-known that the disturbance of immune and inflammatory responses caused by an acute viral infection can lead to long-term disorders. Some examples are the Chikungunya virus, the Ebstein-Barr virus, or other epidemic coronaviruses, such as SARS-CoV and MERS-CoV, which are associated with chronic fatigue, anxiety, and depression [10]. Another relevant issue is the difference between sexes: while men, perhaps due to a previous pro-inflammatory state, are at greater risk in the acute COVID-19, women are more frequently affected in PCS [7]. In this sense, it is believed that the greater specific T-cell response to SARS-CoV-2 in women, may play an important role [11].

Moreover, it has been suggested that after an acute COVID-19 and in predisposed women, a low-grade, continuous inflammatory reaction that increases the risk of PCS would be activated [9,12]. Low-grade inflammation (LGI) is a chronic, systemic, ineffective inflammation that leads to oxidative stress and causes tissue damage [13,14]. The demonstration of higher levels of inflammatory markers in PCS subjects compared to those without PCS could provide evidence of an underlying LGI, and would allow deepening in the relationship between immunity and post-COVID symptoms [11]. However, the available data on biomarkers in PCS are very scarce [7], little systematized, and with contradictory results. Thus, elevation of serum C-reactive protein (CRP) has been reported [4], while other studies did not find any association [15].

Most of the PCS reports have been carried out on post-discharged patients [3], with overrepresented comorbidity and after a moderate or severe COVID-19. In this sense, it has been recommended [16] to study PCS in mild COVID-19, in a community setting, since this approach makes it possible to analyze PCS, reasonably ruling out persistent symptoms due to comorbidities or to organ sequelae.

Establishing the diagnosis of PCS may be difficult for several reasons. Symptoms may appear after a complete recovery from the acute condition and often fluctuate in time and intensity. Moreover, no specific biological marker or sign has been described. Finding a tool that helps in the diagnostic process will facilitate the proper care of these patients.

In light of these considerations, we designed this study to compare the levels of inflammatory biomarkers in subjects with and without PCS after a mild COVID-19 episode. Given the sex differences in the immune response to COVID-19, a secondary objective has been to determine if there were differences in the variables related to PCS development according to gender.

## SUBJECTS AND METHODS

### Participants and sample

This cross-sectional study was carried out on the general population of a semi-urban Basic Health Zone covered by a Primary Health Care center (Camargo-Interior) in Santander, Northern Spain.

The sample was obtained from the medical records of family physicians based on COVID-19 cases between April and September 2020. All cases were followed up exclusively at the Primary Care level and none of them had been vaccinated against SARS-CoV-2. We included confirmed COVID-19 cases by a positive result in the real-time reverse transcription-polymerase chain reaction test (RT-PCR) or by the presence of IgG antibodies against SARS-CoV-2, three months after the acute episode. A second inclusion criterion was a mild course of infection, according to the WHO definition [17], that is, without signs of pneumonia or hypoxia. No exclusion criteria were considered.

The evaluation was carried out 3 months after the onset of the acute episode and consisted of a clinical interview -by phone or face to face- and a blood sample. The interviews were carried out by 3 of the co-authors, using a structured questionnaire and after specific training.

### Study variables

Three groups of variables have been analyzed: epidemiological data, variables from the acute COVID-19 episode, and data at 3 months follow-up.

Sex, age, tobacco consumption, comorbidities -Charlson comorbidity index- and the body mass index - BMI, measured in kg/m^2^-were evaluated. Likewise, disorders related to a worse outcome of acute COVID-19 [18] have been registered, such as immunosuppression (considering lymphoma, AIDS, and active neoplasia under specific treatment), chronic kidney disease, obstructive sleep apnea-hypopnea syndrome (OSAHS),cerebrovascular disease, hypertension, diabetes mellitus, dyslipidemia, ischemic heart disease, chronic obstructive pulmonary disease (COPD) and asthma.

Variables related to the acute episode were the duration (in days), the number, and the following specific symptoms [19]: odynophagia, cough, low-grade fever/fever (>37.5°C), diarrhea, anosmia/dysgeusia, fatigue, myalgia, headache, dyspnea, chest pain, and rhinitis.

Finally, the variables obtained from the evaluation after 3 months of the acute episode (median=115 days), consisted of a structured clinical interview and a blood sample including inflammation markers and SARS-CoV-2 IgG antibodies. Through the interview, the presence or absence of symptoms at that time was collected, and the diagnosis of PCS was established if the criteria of the National Institute for Care and Excellence (NICE) were fulfilled, that is, signs and symptoms that develop during or after an infection consistent with COVID-19, continue for more than 12 weeks, and are not explained by an alternative diagnosis [20].

Blood samples were obtained by the standard antecubital vein puncture procedure in the morning and after 12-hour fasting. Biochemical parameters were analyzed by an automated method on a Technicon Dax autoanalyzer (Technicon Instruments, CO, USA).

### Inflammation markers

Serum CRP, ferritin, lactate dehydrogenase (LDH), fibrinogen, D-dimer levels, and neutrophil and lymphocyte counts have been analyzed. They are routinely used in acute COVID-19 and have demonstrated their usefulness as markers of chronic inflammation [13,21-24]. CRP has been measured in mg/dL, and the detection limit was 0.4 mg/dL. LGI has been defined by a conventionally accepted plasma CRP level between >0.3 mg/dL and <1.0 mg/dL [24]. LDH has been measured in U/L and ferritin in ng/mL, with normal ranges of 120-246 and 22-322, respectively. Fibrinogen was measured in mg/dL, with a normal range between 180 and 500, and D-dimer, in ng/mL, with a normal range between 0-500. Neutrophil and lymphocyte count, with normal ranges between 1.4 and 7.5 (×10^3^/µL), and between 1.2 and 5 (×10^3^/µL), respectively. Likewise, the neutrophil/lymphocyte ratio (NL ratio) has been analyzed. This parameter reflects the intensity of the inflammatory response and the deterioration of the immune system, through the increase in neutrophils and the decrease in lymphocytes, respectively. It is a prognostic marker of severity in acute COVID-19 and has also shown its usefulness as a marker of chronic inflammation [25].

### Statistical analysis

The sample has been analyzed after a double stratification, by sex, and by PCS. After checking the assumption of normality, the quantitative variables have been expressed as mean ± standard deviation (SD) or median [interquartile range (IQR)].

Student’s t and ANOVA tests were used as parametric tests and the median and Kruskal-Wallis tests as non-parametric tests. Categorical variables have been expressed as percentages, and χ^2^ tests (Pearson and Fisher’s exact test) have been used for their comparison. Correlation analyses have been performed using Pearson’s r and Phi coefficient for categorical variables. The quantitative variables have also been transformed and analyzed in a dichotomous way, according to the median of the distributions, considering values lower than the median as a control group. The strength of an association has been expressed as an odds ratio (OR) with its corresponding 95% confidence interval (CI).

To address limitations associated with detecting slight elevations assessing the inflammatory biomarkers separately, composite indices that represent heightened inflammatory activity were built [13]. With this aim, we divided the sample into two random halves and used one of them to build and select the indices. Applying the high ranges of the markers distributions and expressed as criteria, 5 composite indices were finally selected and in a second step, they were applied to the whole sample and to the subsamples of females and males (Table 1). As an example, a case was classified as positive in the D1 index if it presented a neutrophil count equal to or greater than the median (≥3.10 ×10^3^/μL), or a value of the NL ratio in the last tertile (≥1.86).

**Table 1:**
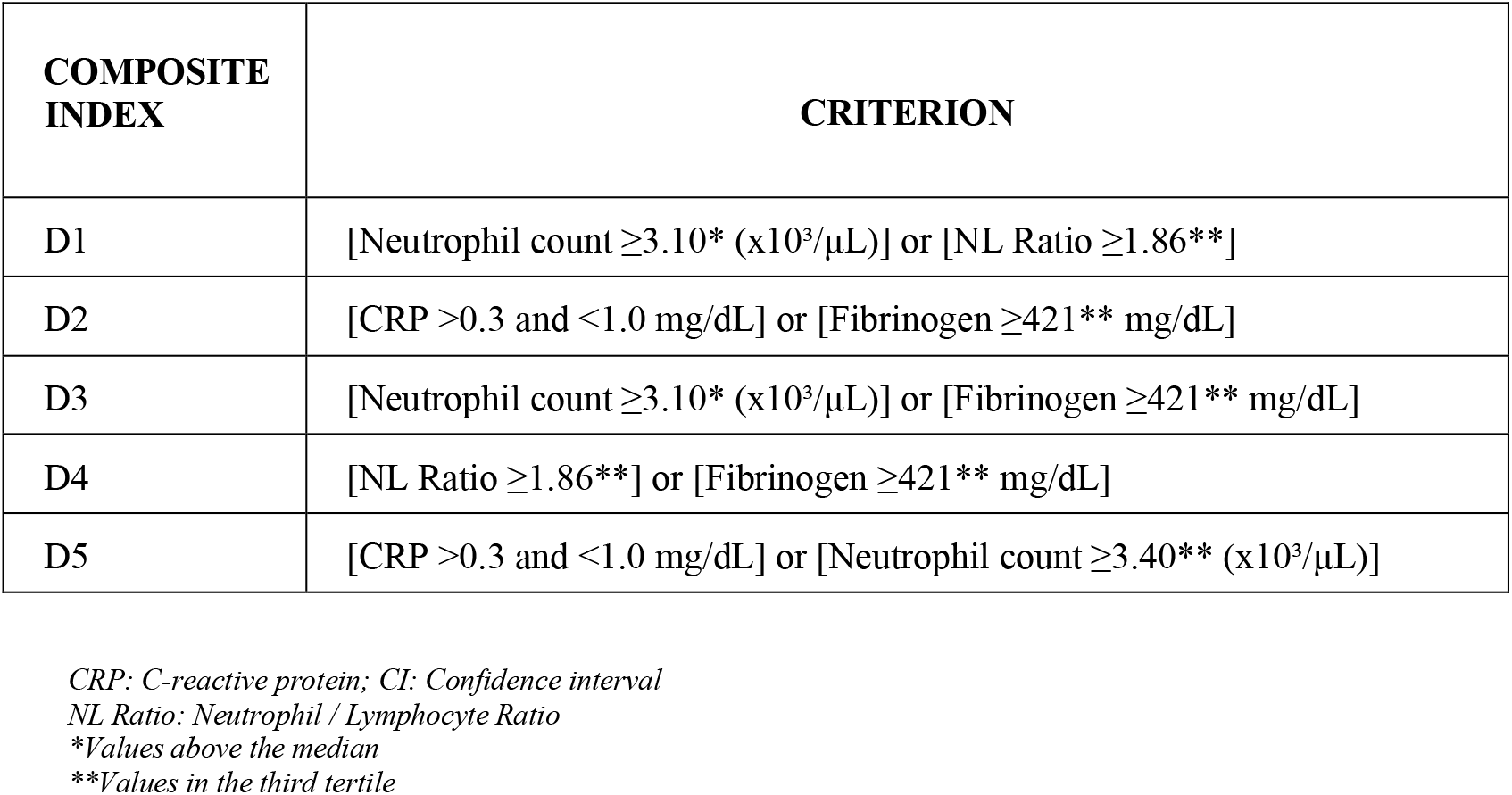
Composite indices of inflammation

| COMPOSITE INDEX | CRITERION |
| --- | --- |
| D1 | [Neutrophil count $\geq 3.10^*$ ($\times 10^3/\mu\text{L}$)] or [NL Ratio $\geq 1.86^{**}$] |
| D2 | [CRP $>0.3$ and $<1.0$ mg/dL] or [Fibrinogen $\geq 421^{**}$ mg/dL] |
| D3 | [Neutrophil count $\geq 3.10^*$ ($\times 10^3/\mu\text{L}$)] or [Fibrinogen $\geq 421^{**}$ mg/dL] |
| D4 | [NL Ratio $\geq 1.86^{**}$] or [Fibrinogen $\geq 421^{**}$ mg/dL] |
| D5 | [CRP $>0.3$ and $<1.0$ mg/dL] or [Neutrophil count $\geq 3.40^{**}$ ($\times 10^3/\mu\text{L}$)] |

CRP: C-reactive protein; CI: Confidence interval
NL Ratio: Neutrophil / Lymphocyte Ratio
\*Values above the median
\*\*Values in the third tertile

Logistic regressions and general linear models, stratified by sex, were performed to ascertain the relationships between inflammation markers and PCS, after controlling for confounding variables. Variables included in the models were: (i) the markers or composite indices that had presented a significant association in the bivariate analysis as independent variables, (ii) the PCS or the specific symptom as outcome variables, and (iii) age, BMI, Charlson index and smoking, as adjustment variables. Logistic regression models were validated by calculating the areas under the ROC (Receiver Operating Characteristic) curves. A value between 0.70 and 0.80 has been considered acceptable, and between 0.80 and 0.90, excellent [26]. The validation of the general linear models was carried out with the analysis of the standardized residuals and their approximation to a standard normal curve. A two-sided p-value <0.05 was considered significant in all the calculations.

### Ethical aspects

Postulates of the Declaration of Helsinki have been carried out. All patients who met the inclusion criteria were informed of the purpose of the study with the delivery of an information sheet and were invited to participate. All of them expressed their verbal consent and there was no refusal to participate. The study was approved by the Cantabria Clinical Research Ethics Committee (Internal Code 2021.102)

## RESULTS

### General characteristics of the sample

Some 212 patients were recruited; the diagnosis of COVID-19 was confirmed in 134, and 13 were discarded for presenting a moderate or severe disease. Finally, the sample consisted of 121 subjects, all of them with mild COVID-19, of which 68 (56.2%) were women. The mean age of the sample was 45.7±16 years, with a range of 18-88 years. Table 2 shows the baseline characteristics of the population and the variables of the acute episode. Overall,the most common symptoms in acute COVID-19 were low-grade fever/fever, anosmia/ageusia, myalgia, and fatigue. Of the acute COVID-19 symptoms, women had a higher frequency of fatigue than men (54.4% vs 30.2%; p=0.008).

**Table 2:** Clinical characteristics of the participants and variables related to the acute COVID-19

|  | **TOTAL**<br>**SAMPLE**<br>**(n=121)** | **WOMEN**<br>**(n=68)** | **MEN**<br>**(n=53)** | ***p*** |
| --- | --- | --- | --- | --- |
| **PERSONAL ANTECEDENTS** |  |  |  |  |
| Age (years) | 45.4 ± 16 | 47.7 ± 15 | 43 ± 17 | 0.12 |
| BMI (kg/m<sup>2</sup>) | 25 [5] | 25 [5] | 25.2 [5] | 0.70 |
| Tobacco; *n*(%) | 43 (36.4) | 22 (33.3) | 21 (40.4) | 0.42 |
| Charlson comorbidity index | 0.4 ± 0.7 | 0.23 ± 0.4 | 0.58 ± 1 | 0.02 |
| Asthma; *n*(%) | 13 (10.7) | 6 (8.7) | 7 (11.3) | 0.62 |
| COPD; *n*(%) | - | - | - | - |
| Immunosuppression; *n*(%) | 3 (2.5) | 2 (3) | 1 (1.9) | 0.70 |
| Chronic kidney disease; *n*(%) | 1 (0.8) | - | 1 (1.9) | 0.44 |
| Cerebrovascular disease; *n*(%) | 1 (0.8) | - | 1 (1.6) | 0.29 |
| Hypertension; *n*(%) | 21 (17.8) | 11 (16.7) | 10 (19.2) | 0.71 |
| Diabetes mellitus; *n*(%) | 6 (5.1) | 1 (1.5) | 5 (9.6) | 0.05 |
| Dyslipidemia; *n*(%) | 38 (32.2) | 23 (34.8) | 15 (28.8) | 0.48 |
| Ischemic heart disease; *n*(%) | 6 (5.1) | 3 (4.5) | 3 (5.8) | 0.54 |
| **ACUTE COVID-19** |  |  |  |  |
| Number of symptoms | 5 [4] | 5 [4] | 4 [4] | 0.50 |
| Duration (days) | 10 [9] | 11 [15] | 10 [9] | 0.40 |
| Cough; *n*(%) | 56 (46.3) | 32 (47.1) | 24 (45.3) | 0.84 |
| Odynophagia; *n*(%) | 39 (32.2) | 23 (33.8) | 16 (30.2) | 0.67 |
| Low-grade fever/fever; *n*(%) | 73 (60.3) | 41 (60.3) | 32 (60.4) | 0.99 |
| Diarrhea; *n*(%) | 18 (14.9) | 11 (16.2) | 7 (13.2) | 0.65 |
| Anosmia/ageusia; *n*(%) | 64 (52.9) | 39 (57.4) | 25 (47.2) | 0.26 |
| Fatigue; *n*(%) | 53 (43.8) | 37 (54.4) | 16 (30.2) | 0.008 |
| Myalgia; *n*(%) | 56 (46.3) | 31 (45.6) | 25 (47.2) | 0.86 |
| Headache; *n*(%) | 51 (42.1) | 28 (41.2) | 23 (43.4) | 0.80 |
| Dyspnea; *n*(%) | 21 (17.4) | 14 (20.6) | 7 (13.2) | 0.28 |
| Chest pain; *n*(%) | 8 (6.6) | 5 (7.4) | 3 (5.7) | 0.71 |
| Rhinitis; *n*(%) | 26 (21.5) | 14 (20.6) | 12 (22.6) | 0.78 |

*Quantitative variables, expressed as mean ± standard deviation or median [interquartile range].*
*BMI: Body mass index; COPD: chronic obstructive pulmonary disease*

### Prevalence and PCS symptoms

Three months after the acute episode, 85 subjects remained asymptomatic and were classified as PCS-, while 36 (29.7%) presented one or more symptoms and were classified as PCS+. The prevalence of PCS in women was 35.8% and in men, 20.8% (p=0.07). Fifteen percent of the PCS+ subjects had ≥3 symptoms, and this subgroup was composed exclusively of women (p= 0.046). Reported symptoms were fatigue (42.8%), anosmia (40%), ageusia (22.8%), dyspnea (17.1%), myalgia (11.4%), palpitations (11.4%), headache (8.5%), telogenic effluvium (8.5%), poor concentration (8.5%), rhinitis (2.8%), cough (2.8%) and nail alterations (2.8%).

Table 3 shows the observed differences in the baseline variables and the acute episode, stratified by sex and PCS. PCS+ women showed a greater number of initial symptoms and a significantly higher frequency of anosmia/ageusia, myalgia, headache, dyspnea, and rhinitis, while in men no difference was noted between PCS+ and PCS-subjects.

**Table 3:** Epidemiological and acute COVID-19 variables in post-COVIDsyndrome

| VARIABLES | WOMEN<br>(n=68) |  |  | MEN<br>(n=53) |  |  |
| --- | --- | --- | --- | --- | --- | --- |
|  | PCS+<br>(n=25) | PCS-<br>(n=43) | *p* | PCS+<br>(n=11) | PCS-<br>(n=42) | *p* |
| Age (years) | 47.2 ± 13 | 47.7 ± 17 | 0.90 | 45.7 ± 17 | 42.3 ± 17 | 0.56 |
| BMI (kg/m<sup>2</sup>) | 23.8 [4] | 25 [5] | 0.31 | 25.1 [5] | 25.2 [4] | 0.94 |
| Tobacco; *n*(%) | 7 (29.2) | 15 (36.6) | 0.54 | 4 (36.4) | 17 (41.5) | 0.76 |
| Charlson comorbidity index | 0.17 ± 0.3 | 0.28 ± 0.5 | 0.29 | 0.27 ± 0.4 | 0.66 ± 1 | 0.27 |
| Number of symptoms | 7 [3] | 3 [5] | 0.0001 | 5 [4] | 4 [4] | 0.37 |
| Duration (days) | 14 [28] | 10 [12] | 0.37 | 10 [8] | 10 [10] | 0.78 |
| Cough; *n*(%) | 12 (50) | 20 (46.5) | 0.78 | 4 (36.4) | 20 (47.6) | 0.50 |
| Odynophagia; *n*(%) | 8 (33.3) | 15 (34.9) | 0.89 | 4 (36.4) | 12 (28.6) | 0.61 |
| Low-grade fever/fever; *n*(%) | 18 (75) | 23 (53.5) | 0.08 | 8 (72.7) | 24 (57.1) | 0.34 |
| Diarrhea; *n*(%) | 2 (8.3) | 9 (20.9) | 0.18 | 1 (9.1) | 6 (14.3) | 0.65 |
| Anosmia/ageusia; *n*(%) | 21 (87.5) | 18 (41.9) | 0.0001 | 7 (63.6) | 18 (42.9) | 0.21 |
| Fatigue; *n*(%) | 17 (70.8) | 20 (46.5) | 0.05 | 4 (36.4) | 12 (28.6) | 0.61 |
| Myalgia; *n*(%) | 16 (66.7) | 15 (34.9) | 0.012 | 7 (63.6) | 18 (42.9) | 0.21 |
| Headache; *n*(%) | 16 (66.7) | 12 (27.9) | 0.002 | 5 (45.5) | 18 (42.9) | 0.87 |
| Dyspnea; *n*(%) | 11 (45.8) | 3 (7) | 0.0001 | 1 (9.1) | 6 (14.3) | 0.65 |
| Chest pain; *n*(%) | 4 (16.7) | 1 (2.3) | 0.05 | - | 3 (7.1) | 0.36 |
| Rhinitis; *n*(%) | 9 (37.5) | 5 (11.6) | 0.013 | 2 (18.2) | 10 (23.8) | 0.69 |

*Quantitative variables. expressed as mean ± standard deviation or median [interquartile range]. BMI: Body Mass Index; PCS+: Post-COVID syndrome; PCS-: No post-COVID syndrome.*

### Inflammation markers

Several significant correlations have been observed according to sex. In PCS+ women, serum CRP was correlated with neutrophil count (Phi=0.51; p=0.015) and with lymphocyte count (Phi=0.43; p= 0.044). In PCS+ men, fibrinogen was correlated with D-dimer levels (r=0.86; p=0.012) and neutrophil count were directly correlated with the NL ratio (r=0.89; p=0.0001), and fibrinogen (r=0.77; p=0.025) and negatively with the lymphocyte count (r = -0.61; p=0.046).

Table 4 shows the crude results of the inflammation markers, including the composite indices of inflammation, stratified by sex and by PCS. In PCS+, compared to PCS-women, a significantly higher prevalence of positive results in the D1, D3, and D4 indices was observed; in the case of PCS+ men, there was a higher prevalence of subjects with a positive result in the D2 and D5 indices, and subjects with CRP in the LGI range (Figure 1). The neutrophil count has shown significantly higher values in women with anosmia, in women with ageusia, and men with fatigue (Figure 2).

**Table 4:**
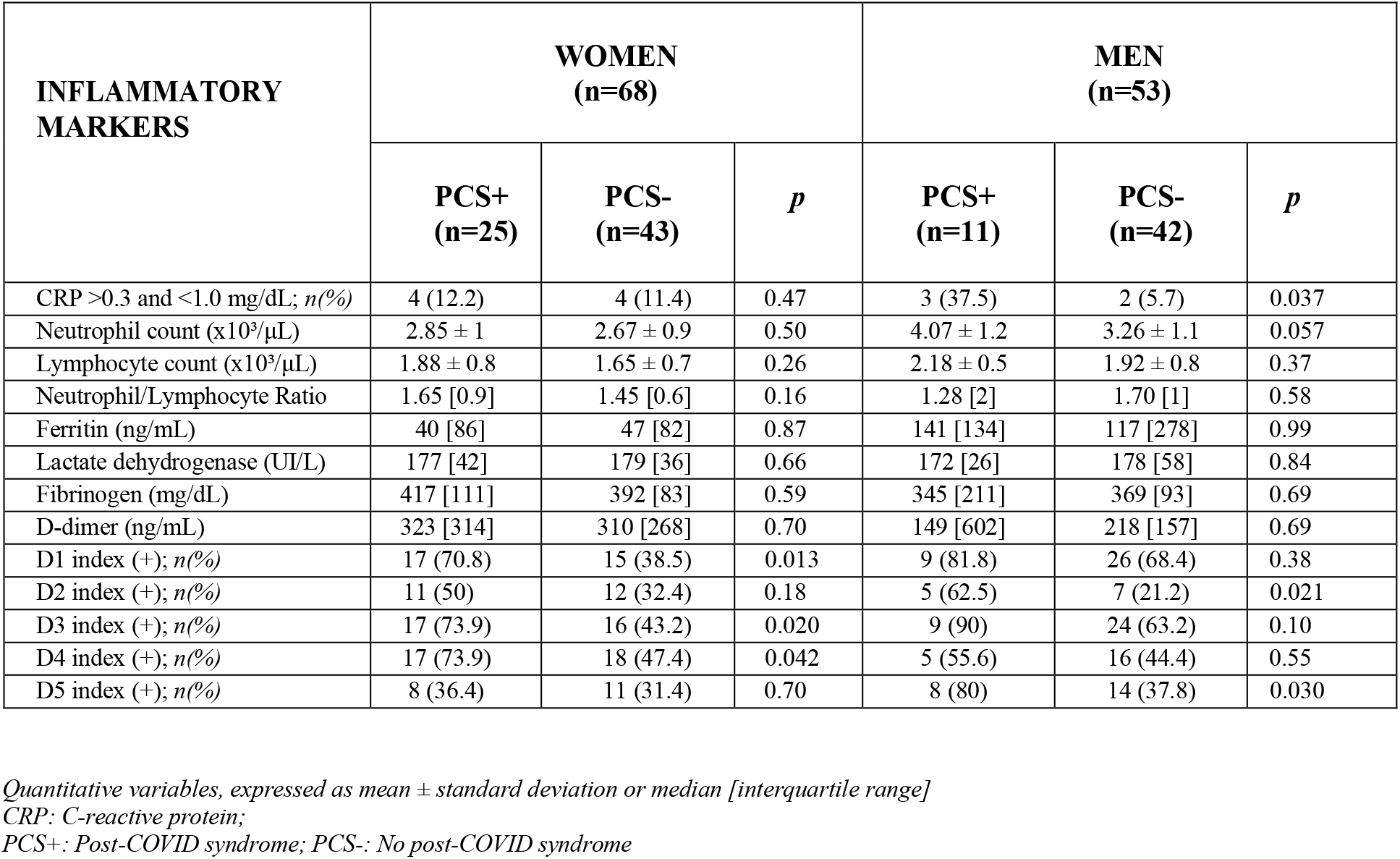
Inflammation markers and post-COVID-19 syndrome

**Figure 1:**
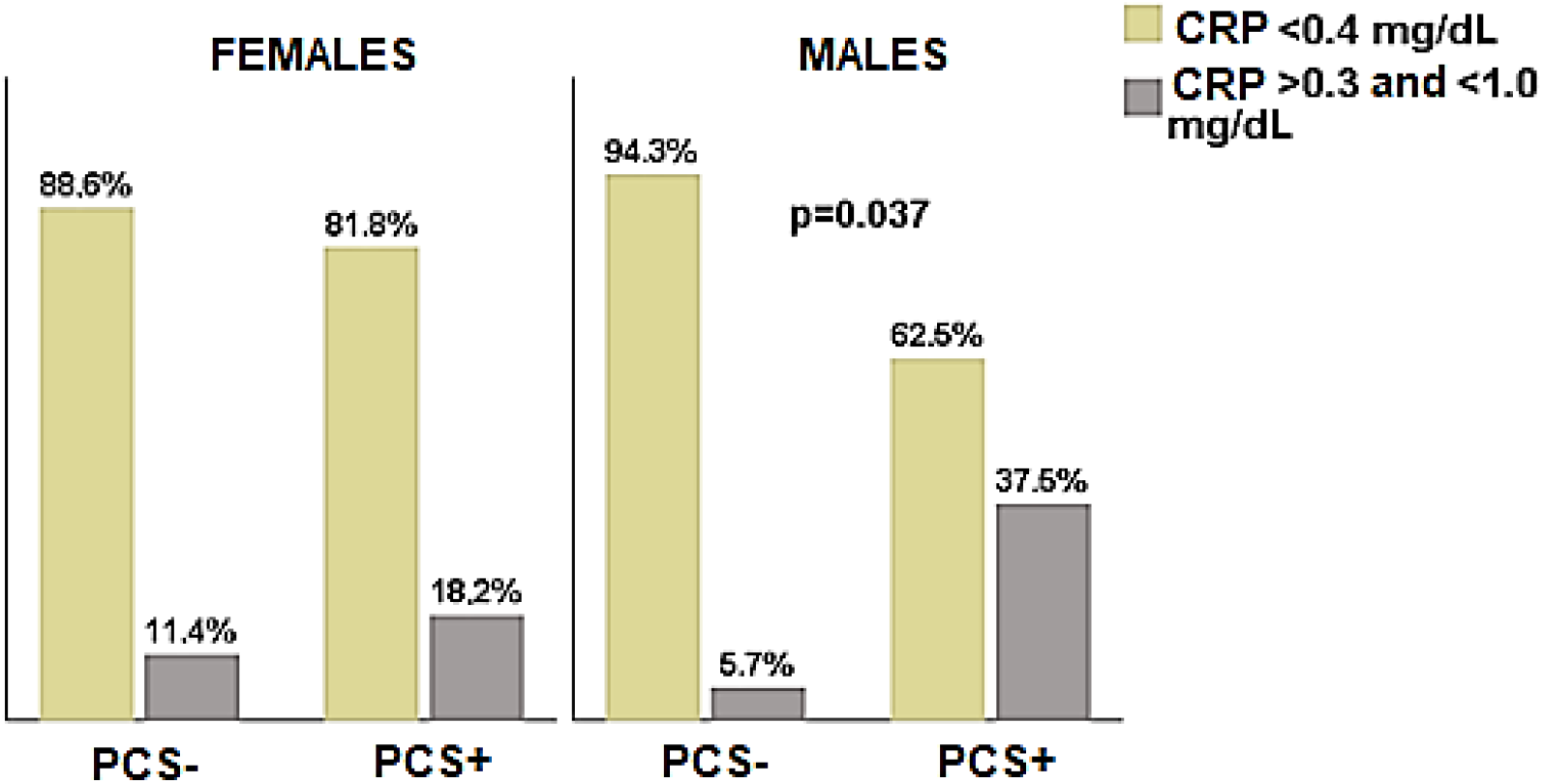
Serum CRP and post-COVID-19 syndrome CRP: C-reactive protein PCS-: No post-COVID syndrome; PCS+: Post-COVID syndrome

**Figure 2:**
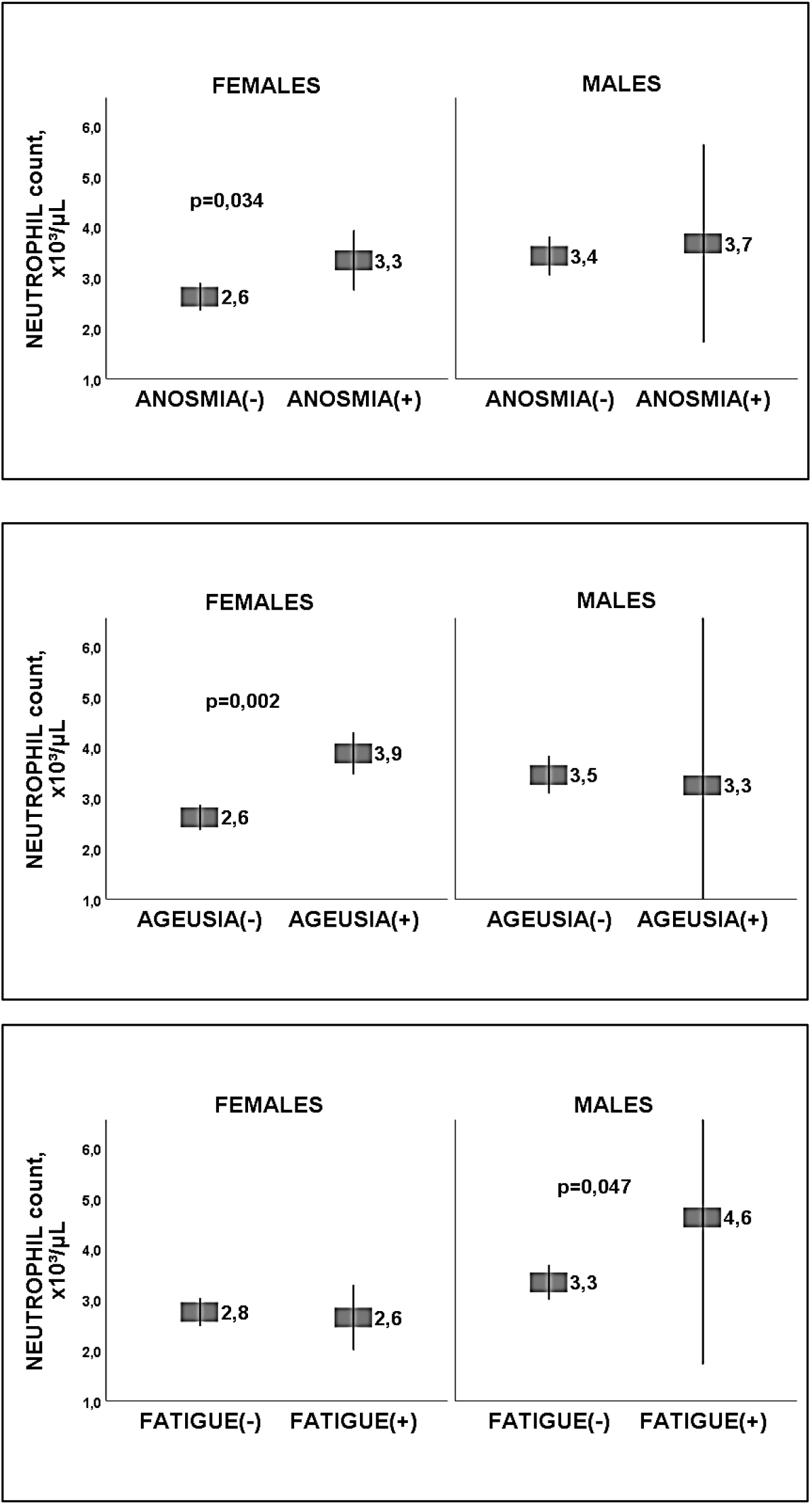
Neutrophil count in anosmia, ageusia, and fatigue, according to gender

In the multivariable analyses, according to the results observed with the composite indices (Tables 1 and 5), and after adjusting for confounders, a woman with a neutrophil count ≥3.10 (×10^3^/μL), with NL ratio ≥1.86 or with fibrinogen levels ≥421 mg/dL, had a 4- to 5-fold increased risk of prevalent PCS. A man with serum CRP >0.3 and <1.0 mg/dL, with neutrophil count ≥3.40 (×10^3^/μL), or with fibrinogen levels ≥421 mg/dL, had a 10- to 17-fold increased risk of prevalent PCS.

**Table 5:** Multivariable models

| INFLAMMATORY MARKER | | Dependent variable | EFFECT SIZE* | | | AUC (CI 95%) / Standardized residuals (mean ± SD) | P (AUC) |
| --- | --- | --- | --- | --- | --- | --- | --- |
| | | | OR (CI 95%) / Estimated marginal means ± SD | p | R<sup>2</sup> | | |
| WOMEN | D1 index | PCS | Adjusted OR=5.14 (1.6-16.4) | 0.006 | 0.27 | 0.76 (0.63-0.89) | 0.001 |
|  | D3 index | PCS | Adjusted OR=4.20 (1.3-13.3) | 0.015 | 0.23 | 0.72 (0.57-0.86) | 0.005 |
|  | D4 index | PCS | Adjusted OR=4.12 (1.3-13.1) | 0.016 | 0.23 | 0.71 (0.57-0.85) | 0.006 |
| | Neutrophil count ($\times 10^3/\mu\text{L}$) | Anosmia | (Anosmia+) 3.43 ± 0.3 (Anosmia-) 2.58 ± 0.1 | 0.014 | 0.20 | 0 ± 0.95 | |
| | Neutrophil count ($\times 10^3/\mu\text{L}$) | Ageusia | (Ageusia+) 3.89 ± 0.3 (Ageusia-) 2.59 ± 0.1 | 0.002 | 0.25 | 0 ± 0.95 | |
| MEN | CRP >0.3 and <1.0 mg/dL | PCS | Adjusted OR=12.9 (1.3-121) | 0.025 | 0.54 | 0.76 (0.58-0.4) | 0.025 |
|  | D2 index | PCS | Adjusted OR=10.1 (1.2-85) | 0.033 | 0.54 | 0.75 (0.55-0.95) | 0.029 |
|  | D5 index | PCS | Adjusted OR=17.5 (2-153) | 0.010 | 0.58 | 0.82 (0.65-0.98) | 0.003 |
| | Neutrophil count ($\times 10^3/\mu\text{L}$) | Fatigue | (Fatigue+) 4.68 ± 0.6 (Fatigue-) 3.37 ± 0.1 | 0.041 | 0.20 | 0 ± 0.93 | |

\* After adjusting for age, BMI, Charlson index, and tobacco consumption.
PCS: Post-COVID syndrome; CRP: C-reactive protein
OR: odds ratio; CI: confidence interval; SD: Standard deviation
AUC: Area under the curve ROC

The AUC values ranged between 0.71 and 0.82, and the standardized residuals showed a normal distribution, with a curve of mean=0 and SD=1 (Table 5). According to these results, the models were suitable for the purposes of the study.

## DISCUSSION

The present study, aimed at analyzing the levels markers of inflammation in PCS, has shown results of interest. Brief mentions can be made on a possible relationship between LGI and PCS, results in the biomarkers, differences by sex, and fatigue and anosmia as the most frequently recorded PCS symptoms.

### Low-grade inflammation and PCS

While acute inflammation is intense, short-lived, and resulting in tissue repair, LGI is persistent, ineffective, and leads to collateral damage [13]. The typical trigger for LGI is damage-associated molecular patterns (DAMP) [13], which are released by damaged cells or tissues as endogenous danger signals, alerting the immune system to non-programmed cell death, invasion by pathogens, and in response to stress [27].

The following sequence can be conjectured: a) COVID-19 infection, b) development of an LGI in predisposed people, c) symptoms with an elevation of biomarkers, d) diagnosis of PCS. It has been observed that the COVID-19 can cause a release of DAMP through different pathways [28,29], potentially contributing to the development of LGI. Besides, frequent symptoms of LGI are chronic fatigue, arthralgia, myalgia, anxiety, depression, constipation, or diarrhea -curiously they overlap with the symptoms of PCS-, and although it does not have specific markers, it is associated with slight elevations of acute-phase reactants and cytokines [14]. In the same line, we have observed slight but significantly higher values of neutrophil count, fibrinogen, and CRP, in patients with PCS or specifically with fatigue, anosmia, or dysgeusia.

Two recently published proteomics studies could corroborate the theory of an underlying LGI in the SPC. Doykov et al. [30] analyzed 96 proteins associated with the immune response in 10 subjects with a positive test for SARS-CoV-2, asymptomatic or after a mild condition, and compared their mass spectrometry profiles with those of similar negative controls. They observed that those who had suffered from COVID-19 had a significant elevation of biomarkers involved in inflammation and the response to stress 40 days after infection, such as the mitochondrial protein PRDX3 or the cytosol protein NDRG1. Holmes et al. [31] analyzed some lipoproteins, glycoproteins, amino acids, biogenic amines, and tryptophan intermediates in plasma samples from patients with past COVID-19 infection (n=27), hospitalized patients with severe COVID-19 respiratory symptoms (n=18), and controls (n=41), using Nuclear Magnetic Resonance spectroscopy and mass spectrometry. Three months after the acute episode, most of these parameters showed a high degree of normalization, but elevated levels of some biomarkers,such as plasma taurine, and the persistence of a reduced glutamine/glutamate ratio were observed, possibly related to a metabolic and inflammatory disturbance.

### Results in inflammation markers: neutrophil count and serum CRP

Neutrophil count has been the most frequently registered marker, with significantly elevated counts in both sexes (in those with fatigue, anosmia, ageusia, and through their participation in the D1, D3, and D5 indices). Figures were located in the upper range of the distribution, without exceeding the upper limit of the normality range, and the differences between PCS+ and PCS-subjects remained significant after adjusting for well-recognized confounders. This result is in line with what is currently known about their complex role in chronic inflammation. Thus, neutrophils are the main effectors of the immune system, it is known that they are a source of DAMP [32] and that in the tissue repair process they can simultaneously release highly immunogenic products that could trigger and/or amplify an inflammatory response [32]. Furthermore, chronic inflammation may in turn stimulate extramedullary neutrophil production and increase their peripheral blood count [33].

On the other hand, serum CRP has been the second most frequent marker, both isolated or included in the D2 and D5 indices. An unexpected finding has been that CRP significant elevations have been noted only in men, but not in women. Serum CRP is an acute-phase protein produced by hepatocytes in response to inflammation, and cytokines such as IL-6 play a key role in stimulating its synthesis [23]. The biological response of pro-inflammatory cytokines during COVID-19 is higher in men than in women [29], and this fact could be reflected in our results.

Evans et al [4] analyzed 1170 patients, discharged from hospital following treatment for COVID-19 and identified 4 clusters of patients suffering from PCS, according to mental and physical impairment. They analyzed plasma CRP according to the clusters and found that patients included in clusters 1 and 2, with very severe and severe impairment, showed prevalences of CRP >1.0 mg/dL of 16.5% and 18%, respectively. On the other hand, people in clusters 3 and 4, with moderate and mild impairment, presented prevalences of CRP >1.0 mg/dL of 11.4% and 6%, respectively. The authors stated that the CRP was particularly elevated in the severe and very severe clusters possibly due to post-COVID-19 systemic inflammation. Unfortunately, there are not available data of plasma CRP with regard to gender.

### PCS and gender

We have observed that PCS affected women more frequently than men, with a trend toward significance, and that the subgroup with ≥3 symptoms was composed exclusively of women. The result is in line with previous studies that systematically indicate that PCS is more frequent in women [4,7,9,34]. Noteworthy, the stratified analysis by sex has also shown some other differences. An example is the association between an acute polysymptomatic episode and PCS. Sudre et al. [34] found that an acute disease with >5 symptoms in the first week was associated with a 4-fold higher risk of developing PCS. Boscolo-Rizzo et al. [35] reported that presenting 3 to 7 initial symptoms was associated with the persistence of symptoms beyond 12 months. Nevertheless, in our study, we have observed that relationship only in females. Thus, an acute COVID-19 with ≥6 symptoms increased the risk of prevalent PCS after adjusting for age, BMI, Charlson index and smoking, with an adjusted OR of 7.1; 95% CI, 1.8-28; p=0.005. Nevertheless, in males, this association could not be demonstrated (adjusted OR=1.06, 95% CI 0.2-4.3; p=0.92).

Another interesting point has been the striking differences in the biomarkers. Some composite indices showed a better performance in women and some others in men. Furthermore, serum CRP in the LGI range was associated with PCS only in men (Table 4 and Figure 1). The explanation probably relies on the different immune responses in both sexes [2]. Women are known to show stronger type 1 IFN responses after stimulation with Toll-like receptor 7 (TLR-7) ligands, have better survival rates than men in certain acute infections, and develop stronger vaccine responses, but also more side effects [2]. Sex steroids, epigenetic factors, and the microbiome -which seems to play a relevant role in modulating the immune response-have also been implicated as etiopathogenic mechanisms of the immune response in women [36].

In acute COVID-19, females have a stronger and higher CD8 T-cell activation response and a lower cytokine and monocyte release [37,38]. While high levels of certain cytokines (TRAIL, IL-15) are associated in women with a worse clinical COVID-19 outcome, poor response of CD8 T cells in men is associated with a worse prognosis [37].

These observations point out to an immunological basis in the different course of the disease according to gender [37], and it could be theorized that these characteristics -the strong CD8 T-cell response, as well as the activation of TLRs and the inflammasome [39]-, are likely involved in the differences by sex observed in PCS.

### Persistent fatigue and anosmia and inflammatory markers

The most frequent symptom of PCS has been fatigue (45.8% of women and 36.4% of men). This high frequency is consistent with previous reports [34,35]. A systematic review of 15 studies targeting PCS (n=47.910), including mild, moderate, and severe cases of COVID-19, concluded that fatigue is the most common symptom [40].

There is an important level of evidence about the role of the activation of peripheral immunity and inflammation in the genesis of persistent fatigue associated with different disorders, such as multiple sclerosis, rheumatoid arthritis, neoplastic diseases, post-viral fatigue, or CFS [41,42].Prolonged post-viral fatigue has been described in Ebstein-Barr, dengue, Zika, Chikungunya, Ross River viruses infection, among others, and has also been associated with other epidemic coronaviruses, such as SARS and MERS [10]. Patients with SARS-CoV and MERS-CoV may present a chronic disease characterized by fatigue, diffuse myalgia, depression, and sleep disorders [43,44] and share profound alterations in the host’s immune system [42]. On the other hand, prolonged post-COVID fatigue has several similarities with the CFS [45]. Thus, both entities have similar symptoms, both can be triggered by a viral infection [46], and the proposed pathophysiological mechanisms are similar -the CFS has been associated with a dysregulation of the immune system, a chronic inflammatory state, oxidative stress, and autoimmunity [47]-.

We observed a higher neutrophil count in men with post-COVID fatigue, a difference that remained significant after adjusting for confounders. The pathophysiological link between neutrophils and fatigue could be the production of reactive oxygen species (ROS) by activated neutrophils, causing oxidative tissue damage. Increased ROS release by neutrophils has been described in situations associated with chronic inflammation, such as age, hyperlipidemia, or hyperglycemia [33]. In contrast, Townsend et al. [15] analyzed 128 people of both sexes (mean age 49.5±15 years; 54% women), after acute COVID-19, and found no relationship between the white blood cell count, neutrophils, lymphocytes, NL ratio, LDH, or CRP, and post-COVID fatigue. The authors analyzed sex as an adjustment variable in the regression model, but they did not perform a differential analysis between men and women.

The second most frequent symptom of PCS was anosmia (41.7% of women and 36.4% of men).. As previously published, COVID-19 anosmia is more common in women [48], young patients [3], and mild forms of the disease [49]. Regarding biomarkers, we found that women with anosmia had a significantly higher neutrophil count. One possible explanation is that young women, compared to men and likely because of the exposure to sex hormones, present an active neutrophil profile characterized by strong type I IFN activity and an increased pro-inflammatory response, which may cause persistent inflammation [50]. Therefore, it could be speculated that the results are related to greater local immunity in young patients, who present mild forms and symptoms of the upper respiratory tract, compared to patients with severe forms, probably older, with lower local immunity, lesser nasal-pharyngeal symptoms, and predominantly lower tract symptoms [51]. In line with this argument, it is worth mentioning that young subjects, compared to older subjects, showed a higher frequency of rhinitis in acute COVID-19 (28.1% of subjects below the median age vs 12.7% of subjects with an age above the median; p=0.040).

PCS has been associated with serum CRP in the range of LGI and with the upper range of distributions of neutrophil count, NL ratio and fibrinogen, with differences regarding to gender. Specifically, in females, a neutrophil count ≥3.10 (×10^3^/μL) and a NL ratio ≥1.86; in males, a neutrophil count ≥3.40 (×10^3^/μL), and in both sexes, a fibrinogen level ≥421 mg/dL. Given that values of biomarkers above these cut-off points could represent a PCS, they might be used as a diagnostic tool in a specific patient. Future research should confirm these results.

The study has several limitations that must be taken into account. Firstly, its cross-sectional design, which allows establishing associations but not inferring causality. It has been carried out on patients from a single Primary Health Care center (semi-urban, in the north of Spain, with a Caucasian population), and the results may not be extrapolated to other populations or geographical areas. Furthermore, successive stratification can lead to a loss of statistical power and a high risk of a type II error.As a point of interest, the composite indices, that have increased the performance in detecting slight elevations of the inflammation markers and at the same time, have provided cut-off points that can be applied in the clinical setting. In addition, the stratified analysis by sex has made it possible to identify differences that would otherwise be hidden, for example, by analyzing sex as an adjustment variable.

## CONCLUSIONS

A consistent association has been observed between PCS after mild COVID-19 and a slight but significant elevation of inflammatory markers. Such a relationship may provide evidence of an underlying LGI in PCS patients.

Neutrophil count was the most frequently recorded marker. The second most frequent marker was serum CRP, and its elevation was only observed in males.

Together with CRP levels between >0.3 and <1.0 mg/dL, the composite indices have provided cut-off points for 3 biomarkers (neutrophil count, NL ratio, and serum fibrinogen). These cut-off points may have clinical relevance as a diagnostic tool and should be confirmed in future research.

Relevant sex differences in PCS and inflammatory markers have been demonstrated, probably related to the different immune responses of women and men to COVID-19. It is mandatory to perform gender-differentiated analyses in the approach to this new and heterogeneous entity.

## Data Availability

The data are available to interested researchers.

---

STROBE Statement—Checklist of items that should be included in reports of **cross-sectional studies**

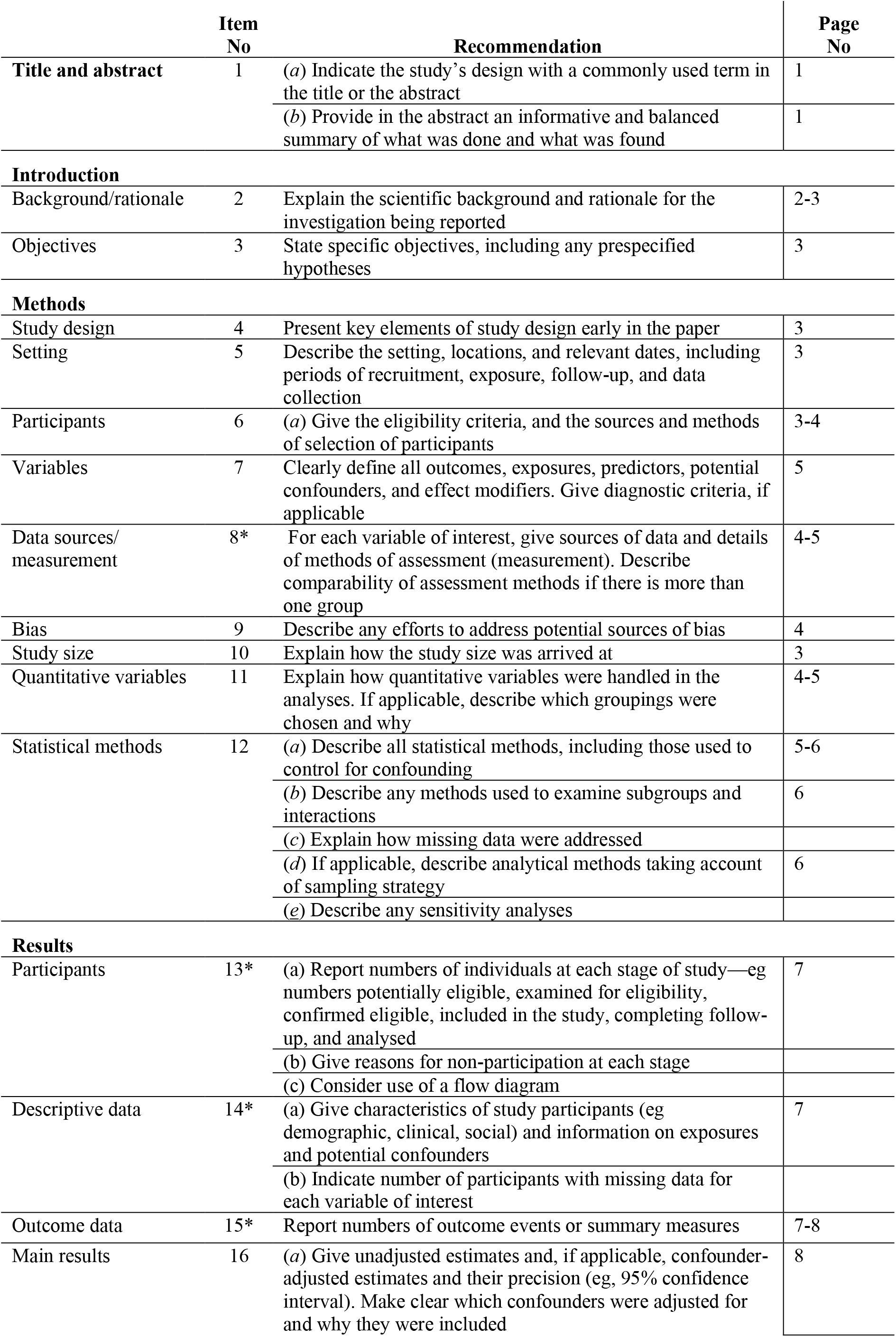
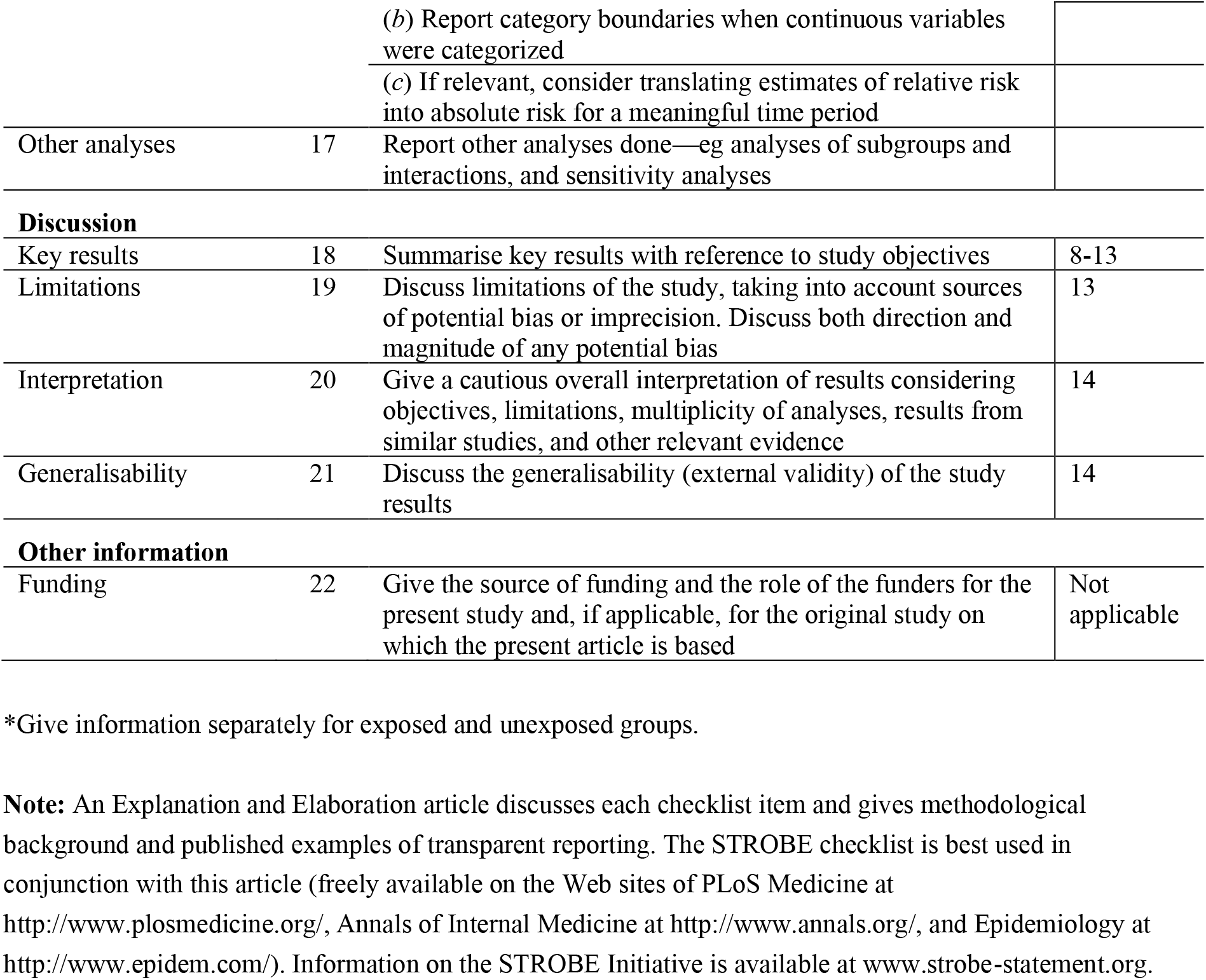

## Notes

**CONFLICT OF INTEREST:** The authors declare that they have no conflict of interest

### Competing Interest Statement

The authors have declared no competing interest.

### Funding Statement

The study has been carried out without any funding

### Author Declarations

The study was approved by the Cantabria Clinical Research Ethics Committee (Internal Code 2021.102)

